# Declining National Codeine Distribution in United States Hospitals and Pharmacies from 2011 to 2019

**DOI:** 10.1101/2022.04.12.22273805

**Authors:** Amy L. Kennalley, Youcef A. Boureghda, Jay G. Ganesh, Adam M. Watkins, Kenneth L. McCall, Brian J. Piper

## Abstract

**Background:** Past research has identified pronounced regional disparities in use of different opioids but less is known for codeine. The primary objective of this study was to analyze the trends of distribution of prescriptions containing codeine in the United States (US) from 2010 to 2019. In addition, this study aimed to identify regional disparities in prescribed milligrams of codeine per person in 2019 and identify any unusual states.

**Methods:** The distribution of codeine via pharmacies, hospitals, and practitioners in kilograms was obtained from the Drug Enforcement Administration’s Automated Reports and Consolidated Ordering System (ARCOS) from 2010 to 2019. In addition, the number of prescriptions of codeine per 1,000 Medicaid enrollees was obtained from the State Drug Utilization Database.

**Results:** The total grams of codeine decreased (−25.0%) through all distributors from 2010 to 2019. The largest increase in total grams of codeine distributed between two consecutive years (2014 to 2015) was +28.9%. For a given distributor type, the largest decrease from 2010 to 2019 was hospitals (−89.6%). In 2019, the total mg of codeine per person distributed in Texas (11.46) was significantly higher relative to the national average (3.06, 1.88 SD). Codeine prescriptions to Medicaid patients peeked in the third quarter of 2016.

**Conclusion:** The peak of prescription codeine in 2011 was consistent with the overall peak in prescription opioids, with a subsequent decrease over the decade. This could be explained by relatively recent recommendations regarding the therapeutic use of codeine and how other antitussive agents may be of better use. The precipitous rise of codeine in Texas that we observed has been recognized in prior studies. These state-level disparities warrant further attention by opioid stewardship committees.

## Introduction

Since 1979, there have been over 600,000 deaths related to drug overdoses in the United States (US) (1). Of those deaths, most have been attributed to opioids, varying from unregulated illegal narcotics such as heroin to prescription opioids (2). When considering that the “first wave” of the opioid epidemic began in the late 1980s, this number offers an even bleaker picture (3). What started as a good-faith effort to be more proactive in managing pain has resulted in over 2 million Americans becoming addicted to some form of opioids (4). Various entities the local, state, and federal levels implemented initiatives directed to combat the problem. One initiative that began in 2016 was The US Surgeon General’s “Turn the Tide Rx’’ campaign, which focused on providing educational resources as well as a voluntary pledge for clinicians to uphold (5). Despite numerous efforts to “turn the tide,” the number of people addicted and dying from opioids continues to increase, so perhaps weaker opioid prescriptions may provide a necessary alternative.

Codeine is a prodrug converted to morphine by the cytochrome P450 enzyme CYP2D6 (6, 7). Morphine, the metabolite that provides analgesic effects, is further metabolized by glucuronidase enzymes to form the inactive metabolite morphine-3-glucoronide and the active metabolite morphine6-glucoronide (6). Genetic polymorphisms of CYP2D6 leads to varied efficacies of codeine based on the phenotypic classifications of ultrarapid, normal, reduced, or poor metabolizer (6, 7). Despite this potentially dangerous variation, codeine is the most prescribed opioid in pediatrics (8).

According to a 2016 Cochrane Review, there is a lack of sufficient evidence to support or oppose the use of cough and cold medications containing codeine in children (9). Various studies have recommended clinicians discover alternatives to codeine for children. This recommendation is due to the variety of effects and the danger of respiratory depression and mortality in children (10, 11, 12, 13). These suggestions are in accordance with the US Food and Drug Administration’s (FDA) 2012 “black box warning” and contraindications for codeine use in children (14, 15, 16).

Depending on the formulation, codeine is classified as a Schedule II, III or V drug in the US (17, 18, 19). The Drug Enforcement Administration’s (DEA) Automation of Reports and Consolidated Orders System (ARCOS) monitors the manufacturing and distribution of all Schedule II drugs as well as Schedule III narcotic and gamma-hydroxybutyric acid (GHB) substances (20). After rescheduling codeine products in Australia to be prescription-only in 2018, researchers examined the drug sales patterns and noted a modest increase (21). Interestingly, another study compared international postoperative opioid prescriptions and found codeine and tramadol accounted for approximately 58% and 45% in Canada and Sweden respectively, and only 7% in the US. However, the US prescribed higher doses of opioids overall following surgery more often than in Canada or Sweden (22). Though the US prescribed comparatively lower postoperative codeine, the number of weaker opioid prescriptions at discharge from the emergency room in the US grew from 2012 to 2017, according to the CDC’s National Health Statistics Report. The formulation acetaminophen-codeine was 12.5% of all opioids prescribed in 2016–2017 (23). To date, there has been no comprehensive study analyzing national codeine prescription trends.

This study’s principal objective was to analyze the distribution trends of codeine-containing prescriptions within the US from 2010 to 2019 based on business type (i.e., hospitals, pharmacies, and health care practitioners). Additionally, an analysis of the distribution of prescriptions containing codeine per Medicaid enrollees from 2014 to 2019 was conducted. Finally, this study aimed to identify regional disparities in prescribed codeine across states and identify any unusual states within the past decade.

## Methods

### Procedures

The ARCOS database, specifically the Retail Drug Summary Reports, was the source of data regarding the national distribution of codeine within the US from 2010 to 2019. The requirement for reporting certain controlled substances transactions from the point of manufacture to distributors — hospitals, retail pharmacies, practitioners, mid-level practitioners, and teaching institutions — to the Attorney General was made possible due to the Controlled Substances Act of 1970. However, since only certain controlled substances are monitored and recorded, it was not possible to consider all the formulations of codeine available nationally. Notably, the Schedule IV and V codeine formulations were not included in the ARCOS analysis (although they are in Medicaid). Report 1 of the ARCOS Retail Drug Summary Reports provided data for grams of codeine distributed by zip code, while the total amount of codeine distributed nationally and by state per quarter (in grams) was collected from Report 4. Additionally, Report 5 was the source for grams of codeine purchased by healthcare business types (i.e., hospitals, pharmacies, and health care practitioners). While Report 5 did contain data on other healthcare business types (e.g., mid-level practitioners and narcotic treatment programs), the amount used by those businesses was negligible. Data on the populations of interest were collected from American Community Survey and US Census Bureau to normalize other findings for a given state or year.

The number of codeine prescriptions covered by state Medicaid agencies reported through Medicaid’s State Drug Utilization Data (SDUD) database from 2014 to 2019 for all 50 states and Washington DC was also collected and corrected for the number of Medicaid enrollees. This study was deemed exempt from review by the Geisinger Institutional Review Board.

### Data-analysis

Values were considered statistically significant via paired t-test with an alpha value of 0.05 or less. Finally, data analysis, figures, and heat maps were completed using GraphPad Prism, Version 9.1.0., JMP, and Microsoft Excel.

## Results

From 2010 to 2019, the total amount of codeine distributed across all the major business types of distributors in the US declined by -25.0% (Figure 1). Further analysis determined that hospitals experienced the largest decrease in codeine distribution with a decline of -89.6%, while pharmacies had a -20.8% decrease in codeine distribution. Practitioners experienced the smallest decrease in codeine distribution of the three business types, with a decrease of -0.15%. Though all business types experienced a decline in codeine distribution in the given period, there was an increase in codeine administration between 2014–2015. More specifically, there was an increase of +28.9% in total codeine distribution across the US (Figure 1). The peak year for codeine distribution was in 2011, with an average of 53.7 mg per person per state. Comparing the peak year to the lowest year in 2019, most states demonstrated a decline in distribution, with an average -40.1% decrease nationally (Figure 2). Figure 2 gives a quantitative illustration of Washington DC and the 46 states which experienced a reduction in codeine distribution over the 10-year focus. In contrast, four states showed an increase in codeine administration: Oklahoma (+28.7%), West Virginia (+35.4%), Arkansas (+28.7%), and Texas (+240.0%) which were all statistically significant (P < 0.05). The heat map also illustrates that Nevada was the state with the most notable decline in codeine distribution between 2011 and 2019, with a significant decrease of - 68% (P < 0.05, Figure 2). Lastly, Figure 3 displays the distribution of codeine (mg/person) in 2019 per state. One state, Arkansas (6.68 mg) distributed a notably large amount of codeine, while four states distributed significantly more codeine (Michigan 6.76 mg, Oklahoma 7.28 mg, West Virginia 8.05 mg, and Texas 11.46 mg) compared to the rest of the US.

**Figure 1.**
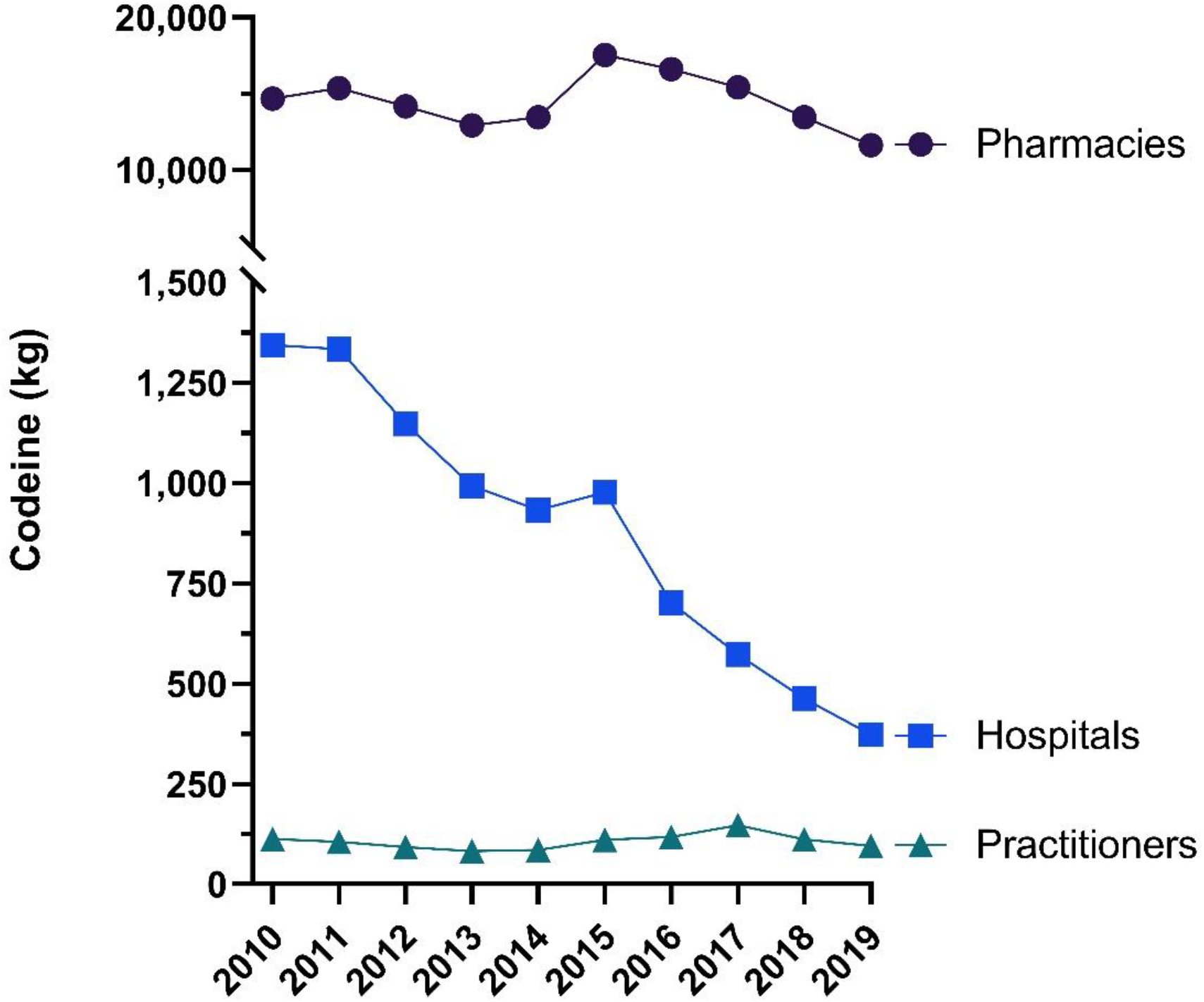
Codeine (kg) by business activity as reported by the United States Drug Enforcement Administration’s Automated Reports and Consolidated Orders System (ARCOS) from 2010 to 2019.

**Figure 2.**
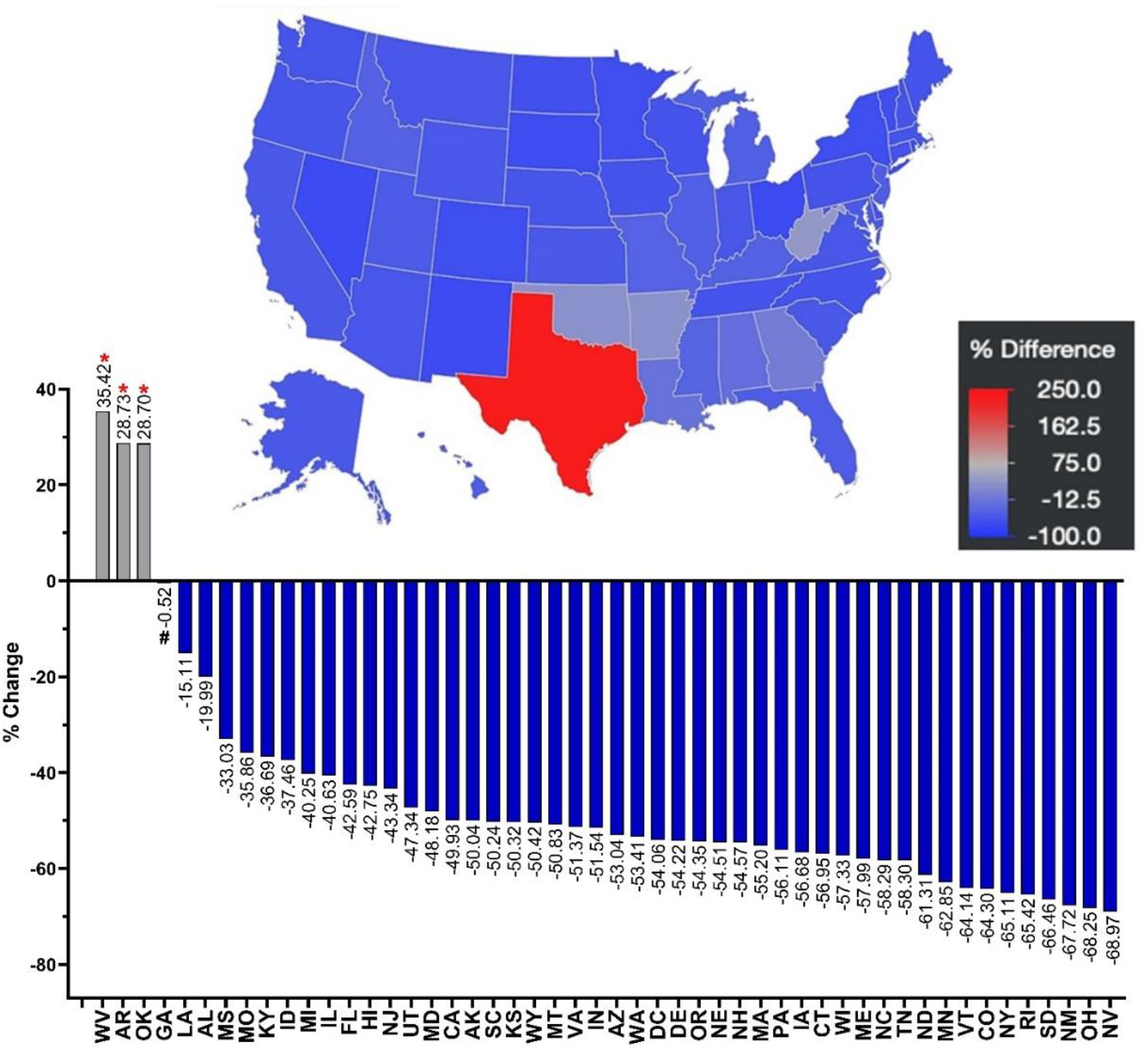
Percent change in distribution of codeine (mg/person) from 2011 to 2019 as reported by the United States Drug Enforcement Administration’s Automated Reports and Consolidated Orders System (ARCOS). Blue indicates a decrease in distribution and red indicates an increase on the heatmap. Bar graph includes 49 states and Washington DC (excluding Texas, +254.0%, for clarity). Percent change between 1.5 SDs and 1.96 SDs from the mean (−45.7%, SD = 23.44%), indicated with a “#”. Percent change > ±1.96 SD from the mean was considered significant (*P < 0.05).

Medicaid prescriptions per 1,000 enrollees were examined between the years of 2014 to 2019. Figure 4 shows that the prescriptions per 1,000 Medicaid enrollees increased to a peak in the third quarter of 2016 with a subsequent drop in 2017, followed by a stabilization to 2019. The total codeine prescriptions in the US increased from the lowest point (21,517 prescriptions) in the first quarter of 2014 to the highest point (391,214 prescriptions) in the third quarter of 2016, reflecting a +1,718.16% increase. Progressing from this peak in the third quarter of 2016 (391,214 prescriptions) to the first quarter in 2017 (166,848 prescriptions) resulted in a -57.35% decrease in prescriptions.

**Figure 4.**
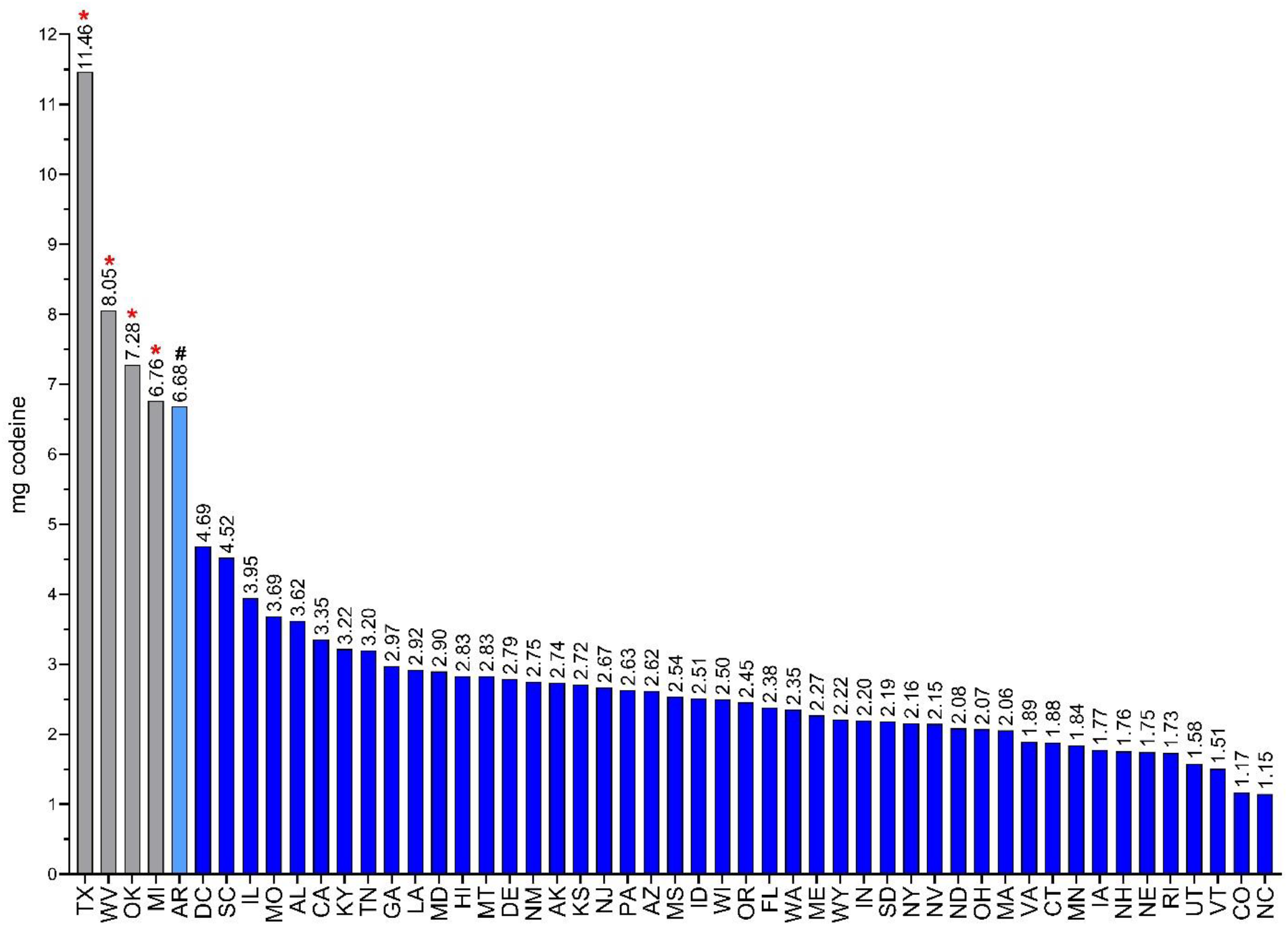
Distribution of codeine (mg/person) in 2019 as reported by the United States Drug Enforcement Administration’s Automated Reports and Consolidated Orders System (ARCOS). Amount of codeine in mg between 1.5 SDs and 1.96 SDs from the mean (3.06 mg, SD = 1.88), indicated with a #. Amount of codeine in mg > ±1.96 SD from the mean was considered significant (*P < 0.05).

**Figure 5.**
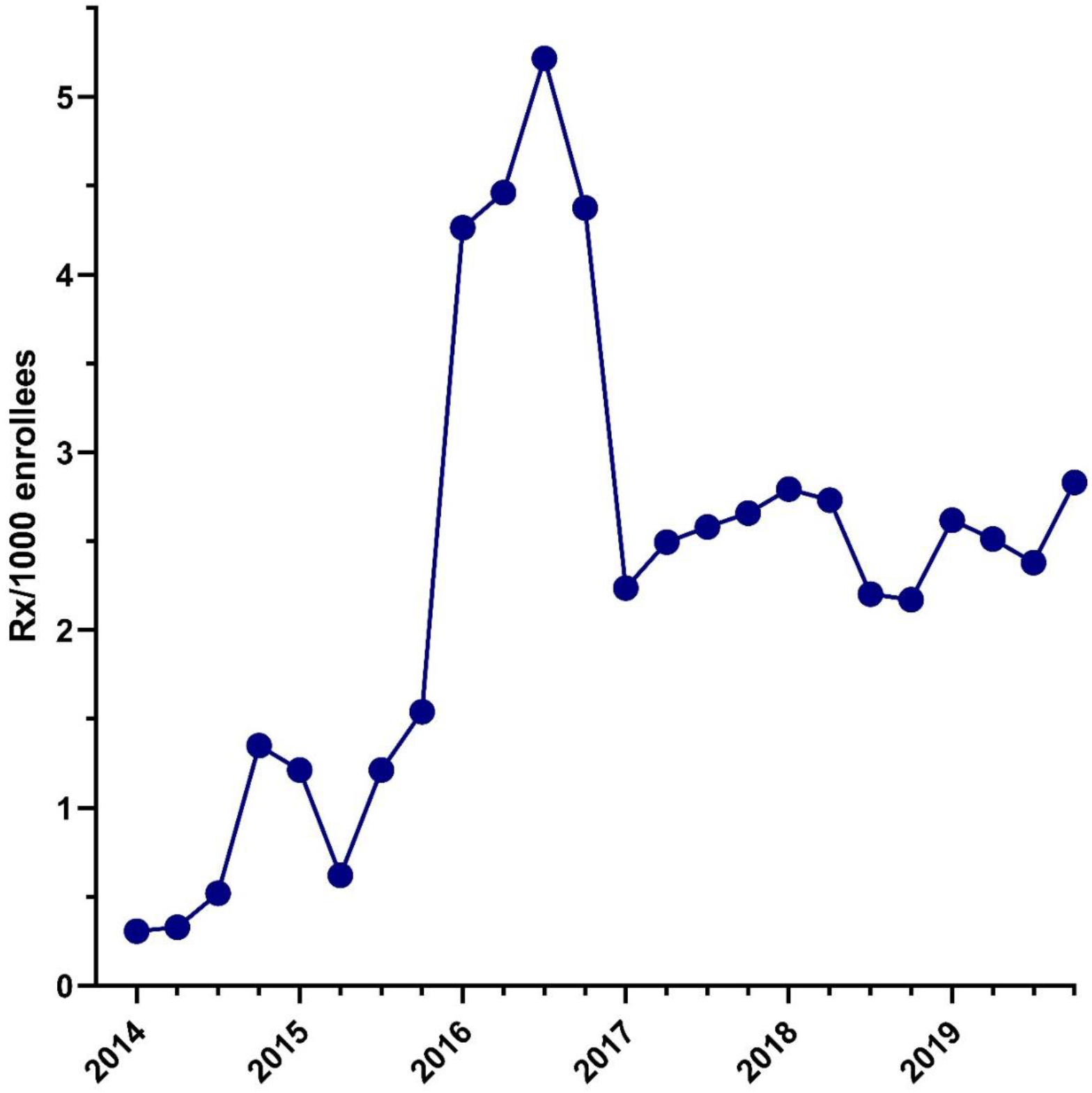
Number of codeine prescriptions per 1,000 Medicaid enrollees as reported by the State Drug Utilization Database (SDUD) from 2014 to 2019.

## Discussion

This study investigated trends in codeine distribution and use in the US between 2011 and 2019, with 2011 corresponding to the peak year for codeine administration. Our data indicates declining distribution of codeine by -25% across all major distributors in the US (hospitals, pharmacies, practitioners) from 2011 to 2019. The change in codeine to outpatients (i.e. distributed to pharmacies, -20.8%) was only one-fourth the reduction to inpatients (i.e. distributed to hospitals, -89.6%). The overall change in codeine nationally is relatively modest relative to those of stronger opioids like hydrocodone, oxycodone, and fentanyl (24, 25). In addition, our data was indicative of regional disparities in codeine utilization when comparing 2011 to 2019. Forty-six out of 50 states experienced a decline in codeine distribution, while four southern states increased from 2011 to 2019: Arkansas (28.7%), Oklahoma (+28.7%), West Virginia (+35.4%), and Texas (+240.0%). Furthermore, an analysis of Medicaid prescriptions per 1,000 enrollees was examined from 2014 to 2019. The data indicated a peak in Medicaid prescriptions during the third quarter of 2016 (391,214 prescriptions per 1,000 enrollees) followed by a subsequent drop in prescriptions during the first quarter of 2017 (166,848 prescriptions per 1,000 enrollees), reflecting a -57.35% decline before stabilizing in 2019.

This pattern of change in the declining distribution of codeine from the peak year of 2011 can be explained with a further examination into other prescription opioids. The peak codeine prescription rate in 2011 was consistent with the overall peak in prescription opioid rates during the same year (20). Comparing the codeine prescription rate between the peak year of 2011 and 2019, the average percent decrease of milligrams of codeine per person was -40.1%. This decrease could be possibly explained by policies that were implemented across the US. As of 2015, 50 states and the District of Columbia put in place an Electronic Prescribing of Controlled Substances (EPCS) system. Moreover, as of April 2021, 24 states have mandatory EPCS protocols in place (26). Given the ongoing trend towards more instituted policies directed towards prescribed substances, such as codeine, this may play an integral role in accounting for opioid prescription reduction. The system’s ubiquitous nature and goal for monitoring the opioid crisis implemented impactful guidelines to control opioid mismanagement.

The US Food and Drug Administration (FDA) guidelines and policies may also be a decisive contributing factor to the progressive decline in codeine distribution. During the period of interest between 2011 and 2019, the FDA instituted policies requiring labeling changes for prescription opioid cough and cold medicine to limit their use to adults 18 years and older (15). Included in these labeling changes is the addition of safety information about the risks of misuse, abuse, addiction, overdose, death, and slowed or difficult breathing in prescription cough and cold medicines containing codeine (16). These mandated FDA policies are likely another contributing factor to the declining distribution of codeine among different healthcare distributors. However, we can also not discount how the general public’s attitudes towards prescription opioids have also changed (27).

According to our regional analysis, 46 of 50 states demonstrated a precipitous decline in codeine utilization when comparing the peak year of 2011 to 2019. However, the four states with an increase in codeine administration were Texas, Oklahoma, West Virginia, and Arkansas. This indicates the presence of a regional disparity in codeine utilizations primarily centered around the southern region of the US in close proximity to Texas, excluding West Virginia. Texas presented with the most notable increase in codeine administration — a +240% increase when comparing 2011 to 2019. According to ARCOS, the spike in codeine distribution for Texas began increasing largely in 2015 and continued to climb with each successive year. The precipitous rise in Texas has been recognized in another prior study (28). A possible explanation for the pronounced regional disparity in Texas, Oklahoma, and Arkansas could be a result from the cultural roots of codeine consumption. Arkansas and Oklahoma also led the US for highest population corrected use of another weak opioid, meperidine (29). On the other hand, the regional disparity could be a result of the complex interplay between variations in health status, attitudes and cultural responses to health care, and access to health care in Texas and its neighboring states. Future research should focus on socioeconomic and policy factors that may mediate the pronounced regional disparity.

Looking to the future, it would be beneficial to further investigate the decreasing trend in codeine in the principal distributors, such as pharmacies and hospitals. Perhaps this decrease resulted from a negative connotation regarding codeine, fear of the drug being misused, or the presence of safer alternatives less likely to cause addiction or abuse such as non-steroidal anti-inflammatory drugs (NSAIDs). Alternatively, it could have been caused by the implementation of stricter guidelines monitoring the control, spread, and prescription of codeine nationally or the adoption of systems designed to distribute codeine more efficiently with the concern about people’s health, safety, and well-being.

Finally, examining the Medicaid codeine prescriptions per 1,000 enrollees, it is clear that further research is necessary to investigate the +1,718.16% increase in prescriptions between the first quarter of 2014 and the third quarter of 2016. According to Medicaid data, the number of national Medicaid enrollees peaked around 2016 and mirrors the rise and fall of prescriptions per 1,000 enrollees between 2014 and 2019. This could be a contributing reason for the increase of codeine prescriptions observed in 2016, but further research should be conducted to discover other possible factors.

In conclusion, gaining insight to combat the opioid epidemic will be aided by further analysis of codeine prescription patterns in the US.

## Data Availability

All data produced are available online at ARCOS Retail Drug Summary Reports.

https://www.deadiversion.usdoj.gov/arcos/retail_drug_summary/index.html

## Acknowledgments

The authors extend their gratitude to Colleen Jordan for her assistance with citations and Iris Johnston for providing access to articles.

## Disclosures

BJP is part of an osteoarthritis research team supported by Pfizer and Eli Lilly. The other authors have no disclosures.

## Notes

### Competing Interest Statement

The authors have declared no competing interest.

### Funding Statement

This study did not receive any funding

## References

1. Jalal H, Buchanich JM, Sinclair DR, Roberts MS, Burke DS. Age and generational patterns of overdose death risk from opioids and other drugs. Nat Med 2020; 26(5): 699–704.

2. Jalal H, Buchanich JM, Roberts MS, Balmert LC, Zhang K, Burke DS. Changing dynamics of the drug overdose epidemic in the United States from 1979 through 2016. Science 2018; 361(6408).

3. Brown R, Morgan A. The opioid epidemic in North America: Implications for Australia. Trends Issues Crime Crim. Justice 2019; (578): 1–15.

4. Kuehn, B. NIH strategy to combat opioid crisis. JAMA 2017; 318(24): 2418–2418.

5. Murthy, VH. Ending the opioid epidemic—a call to action. N Engl J Med 2016; 375(25): 2413–2415.

6. Linares OA, Fudin J, Schiesser WE, Daly Linares AL, Boston RC. CYP2D6 phenotype-specific codeine population pharmacokinetics. J Pain Palliat Care Pharmacother 2015; 29(1): 4–15.

7. Fulton CR, Zang Y, Desta Z, Rosenman MB, Holmes AM, Decker BS, et al. Drug-gene and drug-drug interactions associated with tramadol and codeine therapy in the ingenious trial. Pharmacogenomics 2019; 20(06): 397–408.

8. Cartabuke RS, Tobias JD, Taghon T, Rice J. Current practices regarding codeine administration among pediatricians and pediatric subspecialists. Clin Pediatr 2014; 53(1): 26–30.

9. Gardiner SJ, Chang AB, Marchant JM, Petsky HL. Codeine versus placebo for chronic cough in children. Cochrane Database Syst Rev. 2016; 7(7): CD011914.

10. Rieder MJ, Jong GT. The use of oral opioids to control children’s pain in the post-codeine era. Paediatr Child Health 2021; 26(2): 120–123.

11. Cartabuke RS, Tobias JD, Taghon T, Rice J. Current practices regarding codeine administration among pediatricians and pediatric subspecialists. Clin Pediatr 2014; 53(1): 26–30.

12. Chidambaran V, Senthilkumar Sadhasivam MM. Codeine and opioid metabolism– implications and alternatives for pediatric pain management. Curr Opin Anaesthesiol 2017; 30(3): 349.

13. Kohler JE, Cartmill RS, Kalbfell E, Schumacher J. Continued prescribing of periprocedural codeine and tramadol to children after a black box warning. J Surg Res. 2020; 256:131–135.

14. FDA. Safety review update of codeine use in children; new Boxed Warning and Contraindication on use after tonsillectomy and/or adenoidectomy [Internet]. United States: USFDA; 2013 [cited 2021 May 20]. 4 p. Available from: https://www.fda.gov/media/85072/download

15. FDA. FDA Drug Safety Communication: FDA restricts use of prescription codeine pain and cough medicines and tramadol pain medicines in children; recommends against use in breastfeeding women [Internet]. United States: USFDA; 2017 [cited 2021 May 20]. Available from: https://www.fda.gov/drugs/drug-safety-and-availability/fda-drugsafety-communication-fda-restricts-use-prescriptioncodeine-pain-and-cough-medicines-and

16. FDA. DA Drug Safety Communication: FDA requires labeling changes for prescription opioid cough and cold medicines to limit their use to adults 18 years and older [Internet]. United States: USFDA; 2018 [cited 2021 May 20]. Available from: https://www.fda.gov/drugs/drugsafety-and-availability/fda-drug-safety-communicationfda-requires-labeling-changes-prescription-opioid-coughand-cold

17. US Department of Justice. PART 1308 -Section 1308.12 Schedule II [Internet]. United States: USDEA; 1974 [cited 2021 May 20]. Available from: https://www.deadiversion.usdoj.gov/21cfr/cfr/1308/1308_12.htm

18. US Department of Justice. PART 1308 - Section 1308.13 Schedule III [Internet]. United States: USDEA; 1974 [cited 2021 May 20]. Available from: https://www.deadiversion.usdoj.gov/21cfr/cfr/1308/1308_13.htm

19. US Department of Justice. PART 1308 - Section 1308.15 Schedule V [Internet]. United States: USDEA; 1974 [cited 2021 May 20]. Available from: https://www.deadiversion.usdoj.gov/21cfr/cfr/1308/1308_15.htm

20. US Department of Justice. Automation of Reports and Consolidated Orders System (ARCOS) [Internet]. United States: USDEA; 2021 [cited 2021 May 20]. Available from: https://www.deadiversion.usdoj.gov/arcos/index.html

21. Schaffer AL, Cairns R, Brown JA, Gisev N, Buckley NA, Pearson S. Changes in sales of analgesics to pharmacies after codeine was rescheduled as a prescription only medicine. Med J Aust. 2020; 212(7): 321–327.

22. Ladha KS, Neuman MD, Broms G, Bathell J, Bateman BT, Wijeysundera DN et al. Opioid prescribing after surgery in the United States, Canada, and Sweden. JAMA Netw Open. 2019; 2(9): e1910734.

23. Rui P, Santo L, Ashman JJ. Trends in opioids prescribed at discharge from emergency departments among adults: United States, 2006-2017. Natl Health Stat Report. 2020; (135): 1–12.

24. Collins LK, Pande LJ, Chung DY, Nichols SD, McCall KL, Piper BJ. Trends in the medical supply of fentanyl and fentanyl analogues: United States, 2006 to 2017. Prev Med. 2019 Jun;123:95–100. doi: 10.1016/j.ypmed.2019.02.017.

25. Stemrich RA, Weber JV, McCall KL, Piper BJ. Pronounced declines in dispensed licit fentanyl, but not fentanyl derivatives. Res Social Adm Pharm. doi: 10.1016/j.sapharm.2021.08.001

26. Jameson H. Don’t panic over mandated E-Prescribing of Controlled Substances Laws. Academy of General Dentistry [Internet]. 2020 [cited 2021 May 21]; Newsroom: [about 1p.]. Available from: https://www.agd.org/publications-and-news/newsroom/newsroomlist/2020/08/10/don-t-panic-over-mandated-eprescribing-of-controlled-substances-laws

27. Macy B. Dopesick: Dealers, Doctors and the Drug Company that Addicted America. Little, Brown and Company; 2018.

28. Ighodaro EO, McCall KL, Chung DY, Nichols SD, Piper BJ. Dynamic changes in prescription opioids from 2006 to 2017 in Texas. PeerJ. 2019; 7:e8108.

29. Boyle JM, McCall KL, Nichols SD, Piper BJ. Declines and pronounced regional disparities in meperidine use in the United States. Pharmacol Res Perspect. 2021; 9(4):e00809. doi: 10.1002/prp2.809.

